# Strategic release of lockdowns in a COVID infection model

**DOI:** 10.1101/2020.05.10.20096446

**Authors:** Yi Zhang, Mohit Hota, Sanjiv Kapoor

## Abstract

The COVID-19 Pandemic has impacted the world’s socio-economic system, resulting in a serious health crisis and lockdowns around the world. As the growth of infection slows, decisions regarding easing of lockdown restrictions are required, keeping in view the capacity of the health-care system. In this paper, we quantify the impact of a multi-phased release of the population from lockdown. Using the SIR model for epidemic spread, we design and implement a method to determine the earliest time of release from lockdown restrictions, constrained by a specified threshold on the subsequent peaks of infection. Trade-offs between the threshold and the earliest times of removing lockdown restrictions are illustrated. Additionally, we consider alternative policy decisions where the population is released gradually from lockdown restrictions and illustrate the trade-offs between the rate of release of population and number of active infections.

## 1 Introduction

Decisions regarding quarantine and timing of release of lock-downs have played, and will, play an important role in controlling the spread and impact of the COVID-19 pandemic caused by the SARS-CoV-2 virus [1] with over 3 million cases worldwide at present. The virus is known to be highly infectious, spreads easily, and leads to acute respiratory distress.

A particular concern to policy makers is the prediction of the spread of the virus and the timing of removal of quarantine. The lockdown phase, effectively a quarantine, removes a fraction of the population from contacting the infection. Upon removal of the restriction, the virus would possibly infect the population that emerges from lockdown, starting a new infection spreading phase and resulting in new peaks of infections. Lockdowns have been used in the current pandemic with degrees of success, depending on the fraction of population that respects the lockdown. The lockdown was enforced strictly in China, as well as in Italy and Spain, but these policies were implemented after the infection had spread considerably. While the lockdown has the impact of reducing the infection spread amongst a sizeable fraction of susceptible people, evidence that easing of lockdown restrictions could possibly restart the infection process can be found in recent reports [2]. The need for reducing the stress on the healthcare system [3] requires an analysis of the behavior of the infection spread after the periods of lockdown end. The US has the largest number of active infectious cases and is undergoing considerably stress [4] on the healthcare system. This indicates need for the study of a strategically timed release from lockdown.

In this paper, we study methods to determine the trade-offs between the time of removal of restrictions versus increase in infected cases. We utilize an algorithm that provides the earliest time of removal of restrictions of lockdown while containing the spread of the disease to within a threshold. Two scenarios are considered, one where the restrictions are eliminated in phases and the other where the population is released from lockdown gradually, as will likely occur. We utilize analysis of epidemic spread models, more specifically the SIR [5,6] model (originally from [7-9]). There have been multiple analysis on the COVID-19 pandemic that model its spread including some that account for lack of complete reporting, a problem that further complicates the modeling process [10-18]. Our methods require the initial susceptible population and the infection rates. A regression method is used to fit the model to provide estimates of the parameters of the standard epidemic model. We then use an optimization method to determine the earliest release times for the population under lockdown. The fraction of population under lockdown is estimated using mobility data. Our algorithms to determine the trade-offs utilize a discrete time recurrence to model the behavior of the spread of infection; this models the dynamics of the disease and is also found in the seminal works by Kermack and McKendrick [7-9]. To determine the earliest time of release, our algorithm relies on monotonicity properties of the solution and a logarithmic search over the space of parameters. The results illustrate that sufficiently delayed and controlled release of lockdown is essential. We note that applications to public policy decisions require multiple other factors including demographics. Publicly available data from multiple repositories [19,20] and the states of Illinois and New York are used to illustrate the method.

We summarize our findings: subject to a known fraction of the population in “lockdown”, and determine that:

- If the population on lockdown is to be released in equal sized batches (which is likely), then it appears prudent to wait for a substantial decrease in “active infections”, e.g., to limit the cases to 75% of the peak, wait for the active cases to drop to 50% of the peak before each phase.
- If the population on lockdown is to be released at a steady rate, then that release rate should be quite low. Once the number of active infections drops to a certain level, a 1% rate of release of the lockdown population would still retain the drop in active infections for New York. For Illinois the recovery rate is abnormally slow, an anomaly that could be resolved from gathering more accurate data.
- We show that an adaptive gradual release policy with variable rate, results in maintaining reduction in active infected cases and provides a relatively fast release of the population from lockdown.

The spread of infections alters the behavior of people and the consequent safety measures adopted alter the rate of infection spread. While some current models attempt to capture this [21], our method, which relies on discrete simulation, has the ability to incorporate a time-varying rate of infection. In the current analysis, however, we utilize a constant rate of infection, obtained from parameters derived by fitting the epidemic model to the data.

## 2 Methods

### Models of Pandemic Spread

We investigate both, the SIR and the SEIR models for the states of Illinois and New York, in USA. The models are defined below. Each of these models involve parameters obtained from a regression analysis using available data for each state. The *goodness-of-fit* is evaluated using the values of R^2^ (the co-efficient of determination), and is discussed later in this paper. Our model fitting results in estimating the fraction of susceptible population and utilizing the inverse of the reported recovery rate as a substitute for the infectious period. There is a discrepancy between the typical recovery rate and that estimated by our algorithm pointing to the need for better models to account for a more restrictive infectious period. The focus of this paper is to illustrate the relationship between the growth of infection and release of population from lockdown.

### Standard SIR Model

This model uses three parameters, denoted as - *S*: Size of susceptible population, *I*: Number of infected individuals, *R*: Number of recovered individuals; and assumes that the population size, *N* remains constant.

Changes over time in the population categories are modeled by the differential equations:

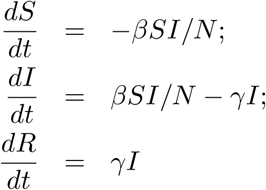

where *β* is the infection rate and γ is the recovery rate.

### SEIR Model

The standard form of the model, without birth and death processes, is modeled by:

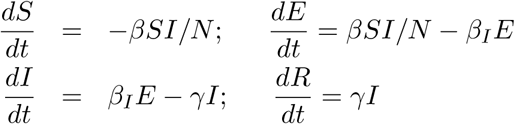

where *E* is the number of individuals exposed to the disease and *β_I_* is the infection rate of the exposed population.

### 2.1 Phased Release Strategy

We use a discrete-step version to compute the model parameters and adapt the above model to incorporate “lockdown”. Let *h* be the fraction of people under lockdown, i.e., the fraction of population that is in “shelter” condition and consequently, (1 − *h*) *S* is the susceptible population. Typically, at the onset of the spread of infection, *h* is a function of time, i.e., *h(t)* which we assume to be a constant for simplicity. Let *l_k_, k* ∈ {1, 2 ... *K*} represent the fraction of population to be released from lockdown restrictions at time *T_k_*, where *K* is the number of lockdown phases. This represents a step function that will model a phased release, corresponding to discrete phases.

#### Optimizing the Release

Our objective is to determine the earliest times of release of the population in phases so as to limit the growth of infected population after the release. The optimization uses the discrete step based mathematical program described below. We assume that all places release the lock down at the same time, allowing us to ignore population changes due to travel. We define indicator variables *Z^k^*(*t*) which are 1 when the *k*^th^ phase of release of the lock down begins. The population enters lockdown at time *t*_0_. The susceptible population is *ηN*, and *h*_0_ is the fraction that is on lock down. It is assumed that the lockdown begins when 200 people are infected. Further, define *E*(*t*) to be the number of exposed to infection population, *I*(*t*) the number of infected people, at time *t*. The modified discrete step SIR model is as follows:

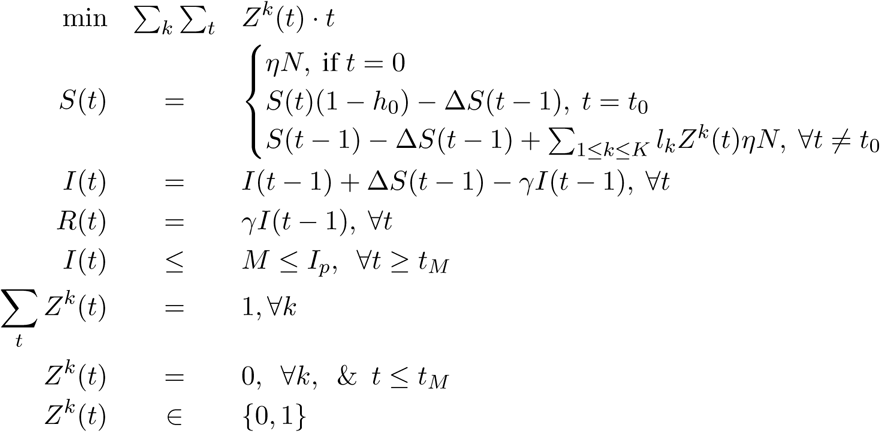

where Δ*S*(*t*) = *βS(t)I*(*t*)/*N* and *M* = TH · *I_P_*; *I_P_* is the number of peak infections under lockdown, and *t_M_* the time at which infections reduce to value *M* after the peak of value *I_P_*, under lockdown conditions and TH is the fraction of the peak that is required to be maintained after lockdown is opened.

Given fixed values of *l_k_* we can solve the above program using a simple search method, resulting in a method that is dependent on the size of the time domain. We next show how to improve this computation.

#### Binary Search for Release Time

Assume the infection reaches its peak at time *t_P_*. Let *S_P_* = *S*(*t_P_*), the number of susceptible people at the time, *t_P_* of peak of active infections, and *I_P_* = *I*(*t_P_*). We get the following

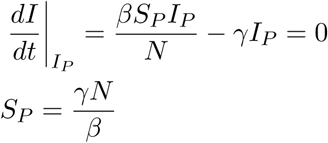

It is clear that at any time *t* > *t_P_*, 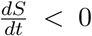 and 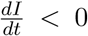. Let *t*_1_*, t*_2_ be 2 different times such that *t_P_* < *t*_1_ < *t*_2_. We consider releasing *F*, a fraction of the lockdown population such that 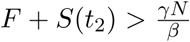 at two different time *t*_1_ and *t*_2_, leading to new peaks 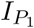 and 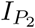 respectively.

##### Lemma 1

*For the SIR model*, 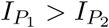

###### Proof

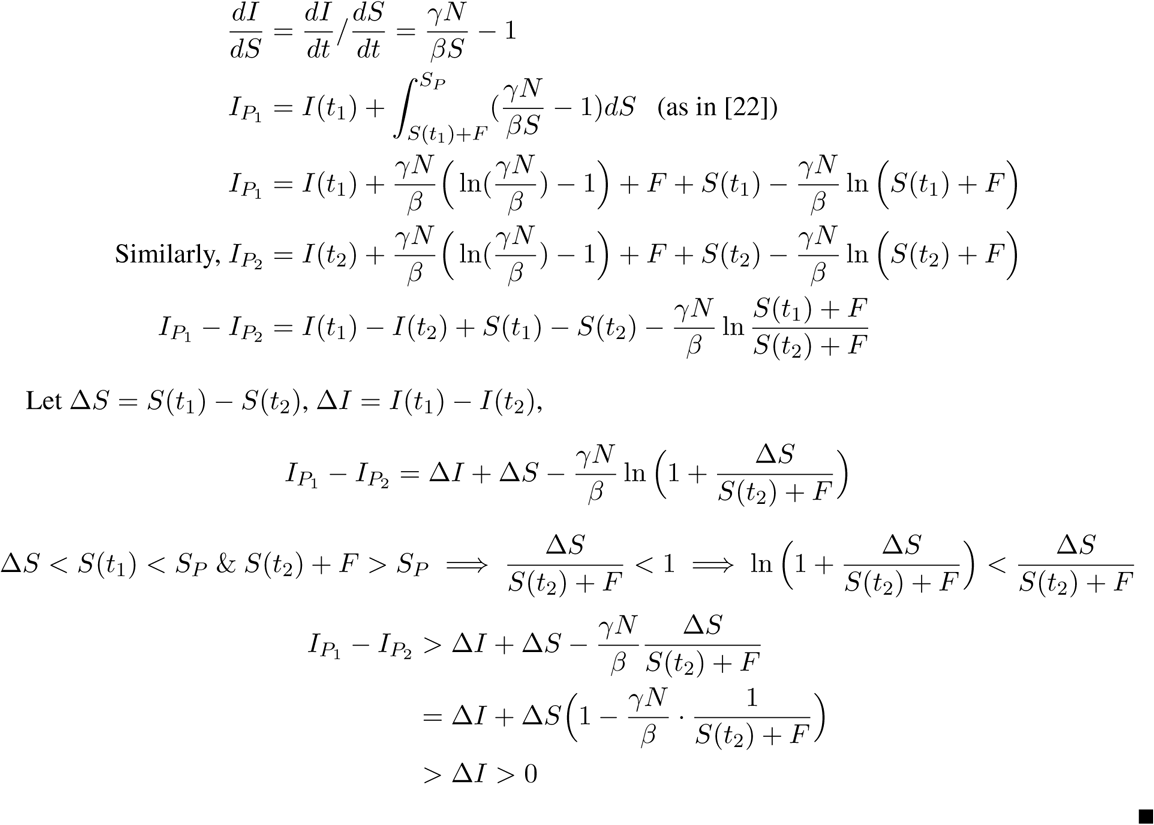

Since the value of the new peak *I_P_* decreases monotonically with respect to the release time, we can perform a binary search to determine the earliest release time that satisfies the peak threshold.

### 2.2 Gradual Release Strategy

Let *δ_S_* be the change in population that we wish to to compute, while maintaining a negative rate of active infections. Changes over time in the population categories is expressed as:

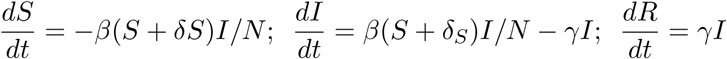

where *β* is the infection rate and *γ* the recovery rate. To maintain the rate of infection decrease to be *r_a_* < 0 we obtain:

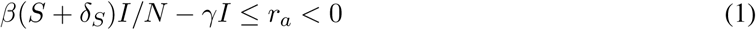

or

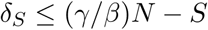

## 3 Results and Discussion

We investigatethe trade-offs between the time of release of lockdown and the consequent impact on the number of infections. In the analysis, a fraction of the population, *h*, that is under lockdown is eliminated from consideration during the initial stages of the infection, and is then re-introduced into the model when the lockdown restrictions are relaxed. Removal of restrictions of the lockdown impacts the susceptible population. We consider two strategies for removal of restrictions. In the first strategy restrictions are relaxed on an equal fraction of the population at distinct times and we determine the earliest time of such release, subject to conditions on growth of active cases. In the second scenario the population is released gradually in small fractions over time and we determined the impact of the release strategy on the number of active cases. We focus on the number of active cases as this metric is important for health-care systems.

The current population under lockdown is estimated using the mobility numbers to roughly 50% average (The Google COVID-19 Community Mobility Reports [23] indicate a 45% decrease in transit and workplace mobility in New York). The analysis of the spread of the infection utilized both the SIR and SEIR models, and in order to fit the models we minimized an error function. The parameter space includes the size of the initial susceptible population, *β* and *γ*, the parameter for recovery time, guided by reports that the typical recovery time have a mean value of 14 days [24]. However the recovery data did not indicate a mean recovery period of 14 days but a much longer period. We describe the determination of the model parameter in a later section 4. Details of these parameter estimates are provided for two states, Illinois and New York. The SIR model provided a more accurate fit of the model and is used in our results on release strategies.

### 3.1 Phased Removal of Restrictions

In the analysis of a phased release, a fraction of the population, *h*, that is under lockdown is eliminated from the population in the system, and this fraction is re-introduced at later stages. Given the model parameters, we introduce this population in phases, releasing a third of this population at every phase. Other release proportions are allowed to be easily incorporated. This population cannot be introduced before the peak of infection, measured by the number of active cases and is illustrated in Figure 1 where the active cases grow substantially when a fraction of the population is released from the lockdown just around or before the peak. In both the states, gradual reopening at a rate of 1.5% of the population under lockdown results in a spike of cases (Figure 1), with Illinois projected to be worse. We note that the data for recovered cases in Illinois is sparse and impacted the projections adversely. The two projections illustrated are for two scenarios (a) 2 weeks after the number of new cases peak and (b) 2 weeks after the peak of the active infected cases.

**Figure 1:**
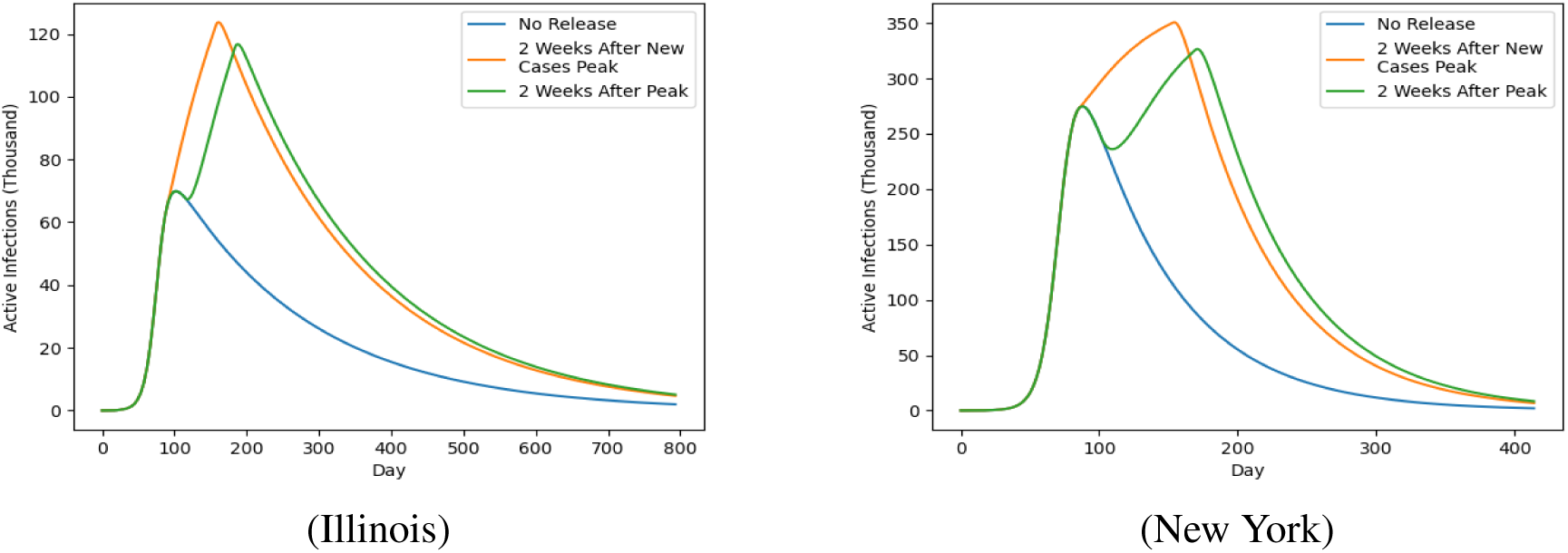
Release prior to peak of active infections (population under lockdown released gradually at a rate of 1.5%).

#### Optimizing the Release

The SIR model is used to compute the peak of active infections. We note that the parameters also include the susceptible population size, which is required to be estimated. We account for the “lockdown effect” by decreasing the susceptible population since lockdown effectively removes a sizable fraction of the population from the infection dynamics. This hidden population is returned to the susceptible population in 3 phases, uniformly, and at time instances that are computed by the algorithm. We set a bound on the projected future growth of the active infections, as provided by the SIR model. This bound TH is set to be a percentage of *I_P_*, the number of active infected cases at the peak of the spread of the virus.

The objective of our algorithm is determine *T*_1_, *T*_2_ and *T*_3_, i.e., the earliest times at which restrictions are removed under the constraint that the active cases do not exceed the threshold percentage (TH) of the number of active infections at the peak that occurred during lockdown. Our optimization method has been detailed in section 2.1. For the states of New York and Illinois, the earliest times of release of each of the three phases and the consequent behavior of the release of the hidden population is illustrated in Figure 2 and Figure 3. Releasing population in phases will result in increase in number of infections and the increase in the active cases is illustrated in Figure 2, while the rise and the pattern of growth in the cumulative infected cases is shown in Figure 3. In this simulation, it is assumed that 50% of the population in Illinois and New York is under “lockdown” after the number of infections hit a figure of 200. This half of the population is released from lockdown in equal phases. The figures do not account for additional measures that may be used against the spread of the infection, including testing and stricter social distancing in public transport and public places. These measures will alter the model parameters.

**Figure 2:**
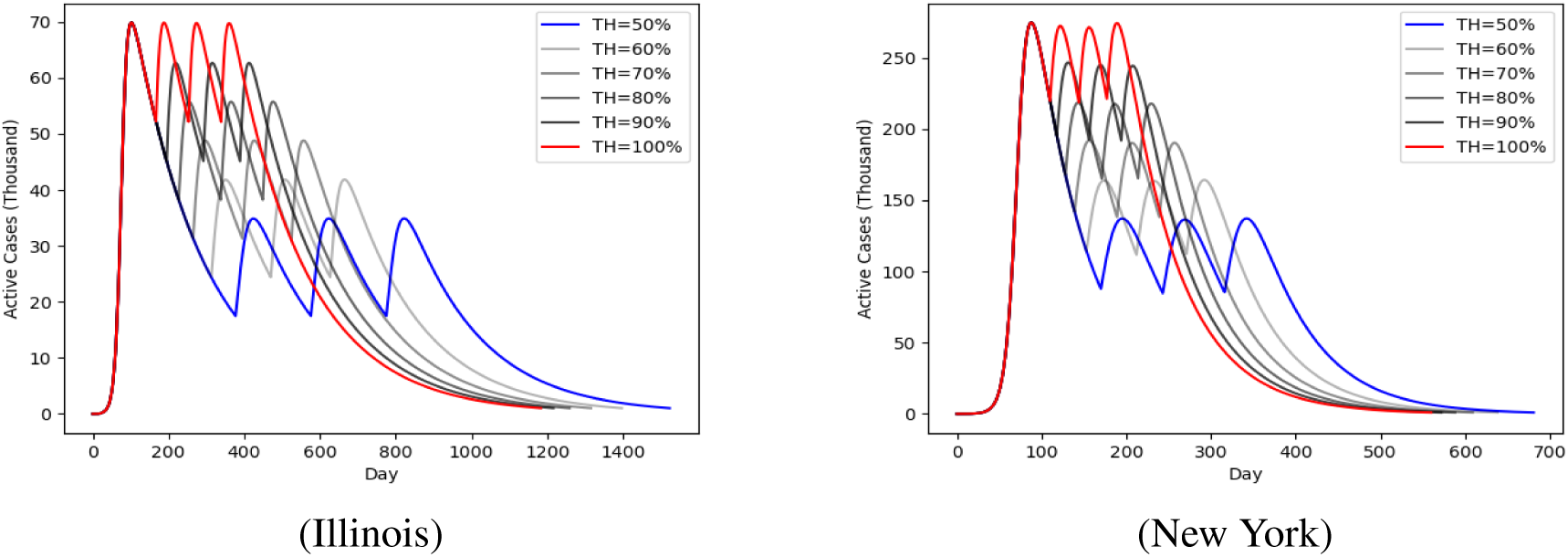
Active infections as a function of days for different threshold percentage TH during a three phase release plan.

**Figure 3:**
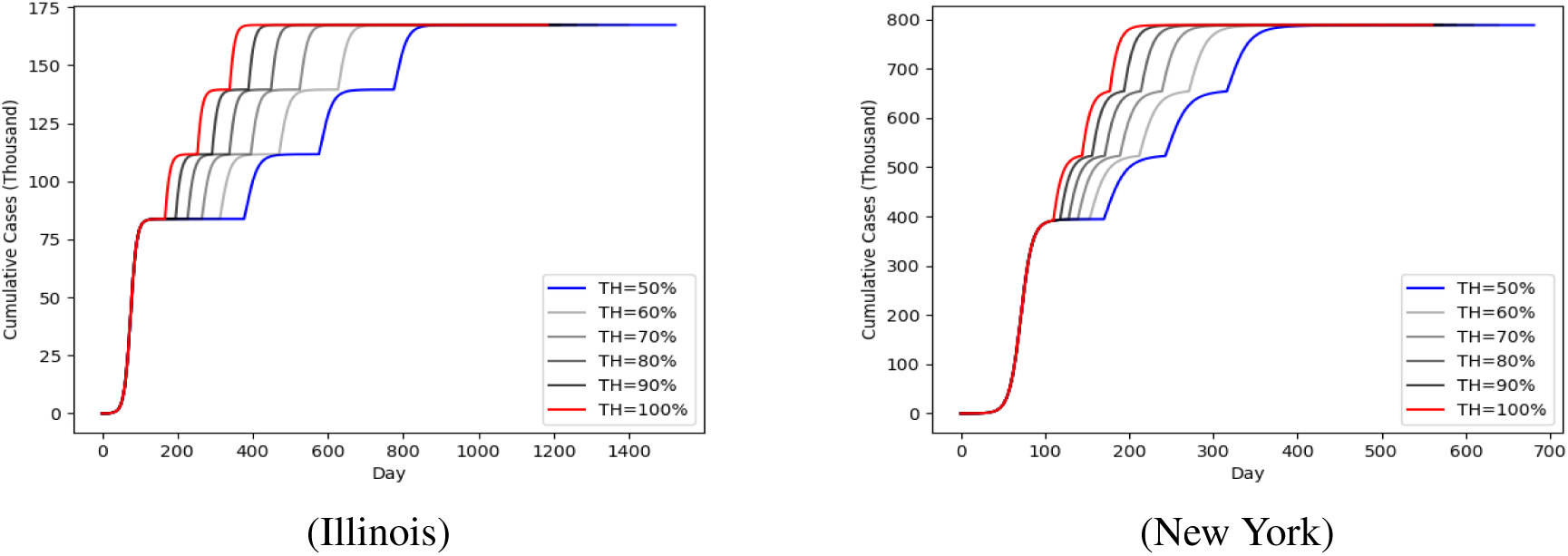
Cumulative Infections as a function of days for different TH, peak threshold during a three phase release plan that limits infection to threshold percentage TH.

#### Impact of Infection Transmission rates

The estimate of the transmission rate is dependent on a number of parameters and may vary. We consider the variation in this factor in determining a relationship between the threshold percentage, TH, and the percentage drop from the peak at which the first hidden population is to be released from lockdown (Figure 4) when optimizing for the release time. This relationship is approximately linear. The measure of infection rate, *β* is used as a variable to compute this relationship for the range of *β* ∈ [*β* * .6, *β* * 1.4], given the current estimate of *β*. This relationship can be used to determine the lockdown release expressed as a percentage of the peak of active infections, given a threshold percentage TH of the first peak of active infection cases that can be afforded by the health-care system without stress. As an example, if the health-care system has the capacity to handle 75% of the first peak, then the release of the lockdown should be at around 50 percent drop from the peak active cases in Illinois. For New York, the corresponding drop is estimated at 50%.

**Figure 4:**
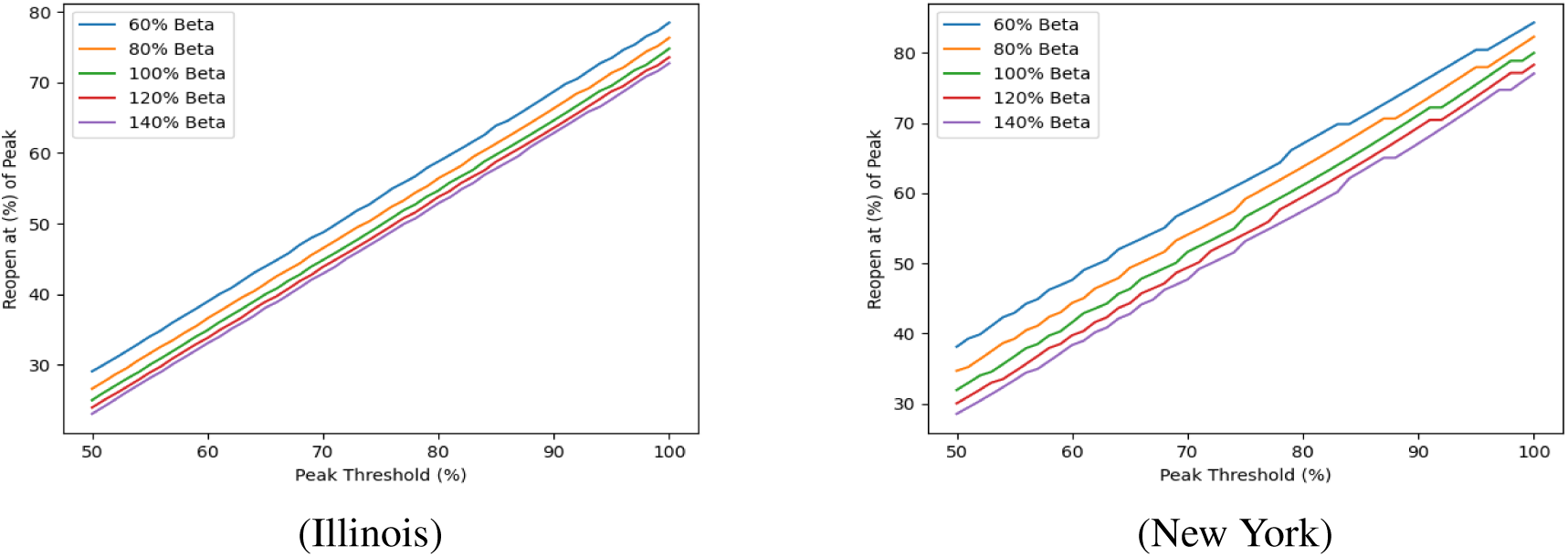
Percentage drop from peak infection required for reopening vs threshold percentage TH.

In terms of number of days till removal of restrictions, Figure 5 illustrates the relationship between the earliest release day of the first phase, as a percentage of the day to the peak infection rate versus threshold percentage, TH, that the active infected cases are to be limited to. As an example, in Illinois, the relationship shows that to limit further infections to TH value of 75% of the active infection peak cases under lockdown, reopening the economy should start after waiting for at least 1.5 times the number of days to the first peak. This is more applicable to counties which are most affected by the spread of infection.

**Figure 5:**
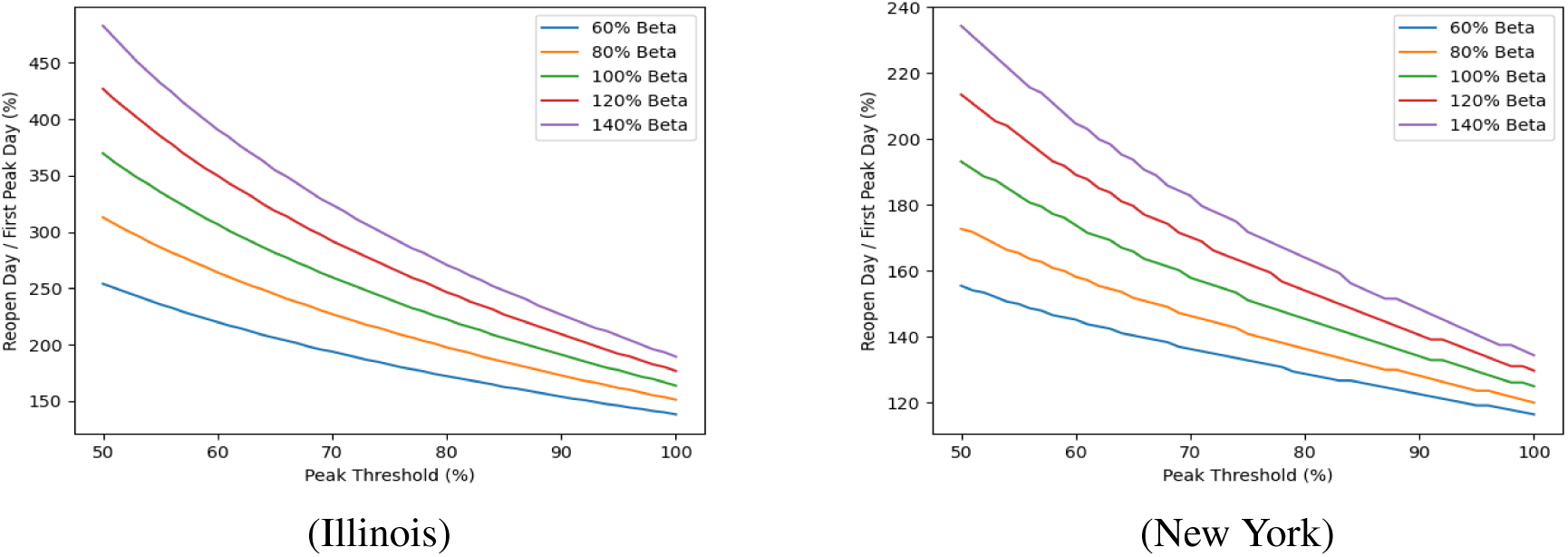
Number of days to restart / Number of days to peak vs threshold percentage TH.

### 3.2 Gradual Release of Restrictions Policy

We contrast the phased removal of restrictions with a graded release policy. The first gradual release policy considered is as follows: A percentage, *κ*, of the original population under lockdown is released linearly, starting at 14 days after the peak. The results are illustrated in Figure 6 for various values of *κ*. The results show that the release should be very gradual.

**Figure 6:**
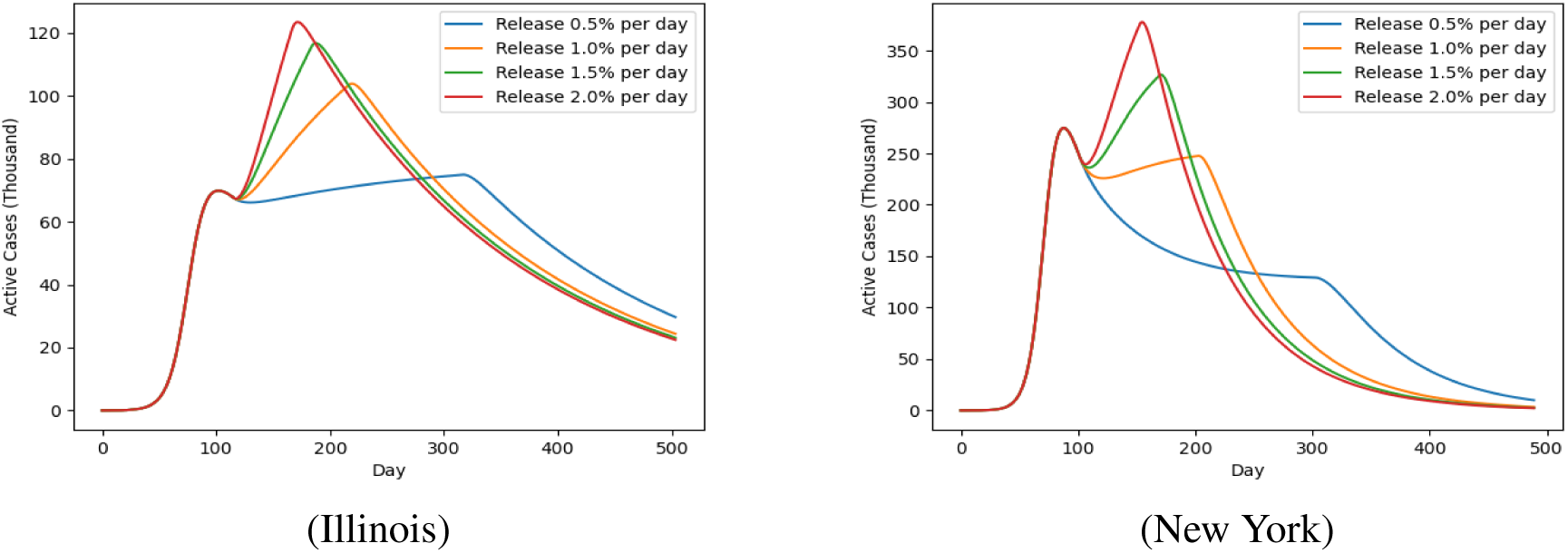
Number of active infections for various linear release percentages. The percentage of population under lockdown is assumed to be 50%.

#### Gradual Policy to uniformly reduce active infections

One possible policy would be to release a percentage of the population with the objective that the number of infections do not increase. Let *δ_S_* be the increase in population and *r_a_* the rate of decrease of active infections. In order to ensure that *r_a_* is negative (see inequality (1)), the relationship

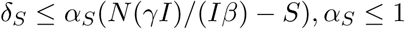

must hold and we utilize this relationship to determine the percentage of population from which the lockdown restrictions are removed. In our simulations, we choose different values of *α_S_* where *δ_S_* = *α_S_*((*γ*/*β*)*N* − *S*), with *α_S_* regulating the release of population as follows: When *α_S_* = 0 no new population element is released, while *α_S_* = 1 indicates releasing a portion of the population that results in no decrease of active cases. Note that this release rate is a function of time. The graph of the active infections (Figure 7) for various release factors shows that the efficacy of the gradual release policy.

**Figure 7:**
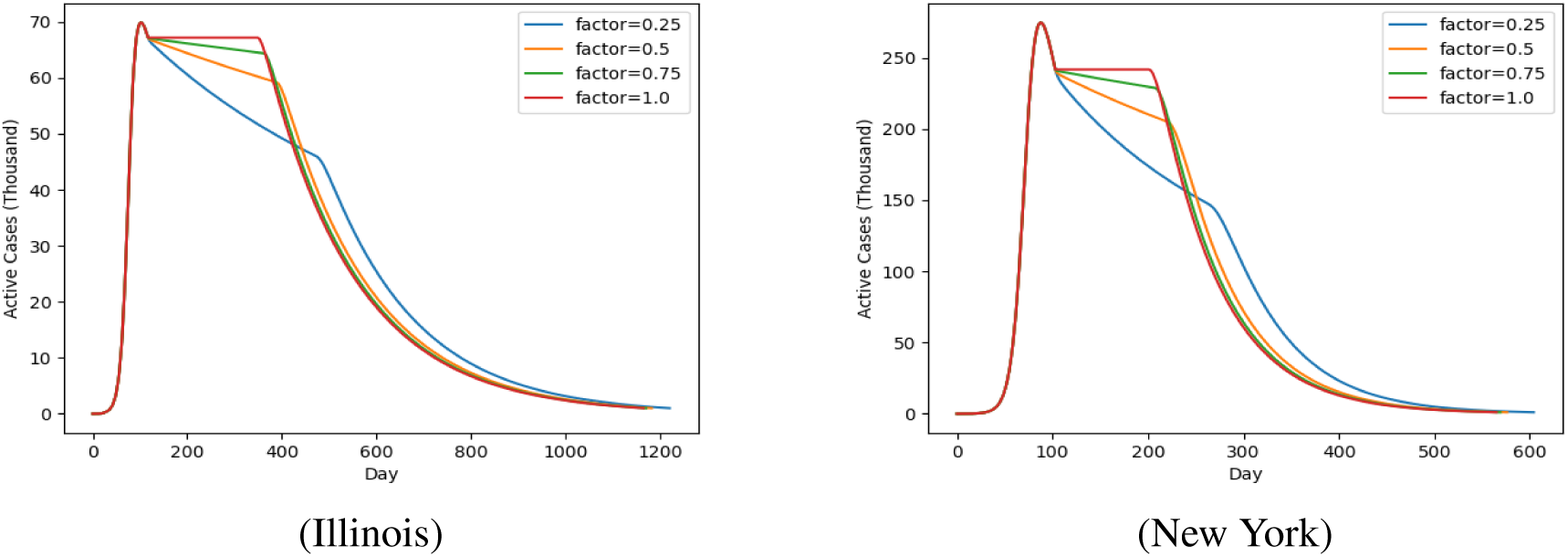
Active Infections vs lockdown release factor, *α_S_*

A measure of efficiency of this mechanism is the days required to complete the removal of restriction, termed “Release-end”. The time of start of this release policy also impacts the “Release-end” days. The gradual release policy is to be activated at the peak of active infections and to start after “delay” days. The decrease relationship between the Release-end days and the release factor *r_I_* is illustrated in Figure 8 for various values of “delay”. Figure 9 illustrates the increase relationship between the Release-end days and the delay for various release factors.

**Figure 8:**
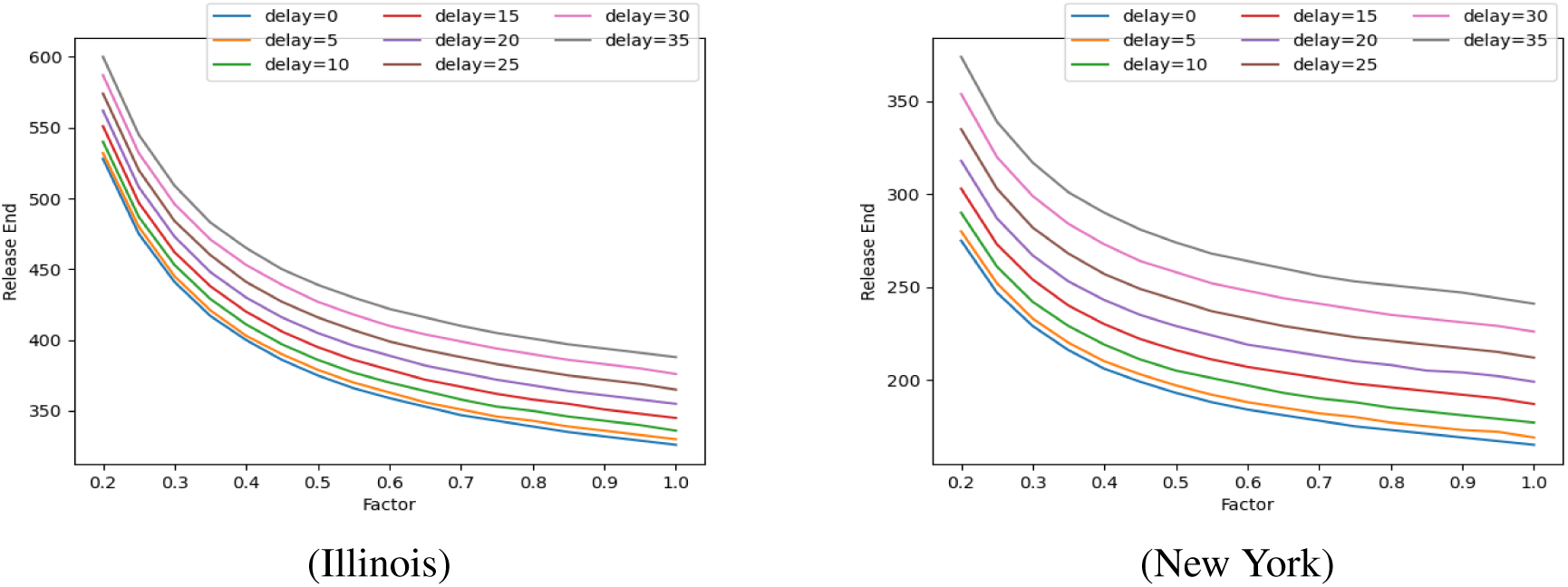
The number of days required for complete removal of lockdown vs rate of release from lockdown, *α_S_* for different start delays.

**Figure 9:**
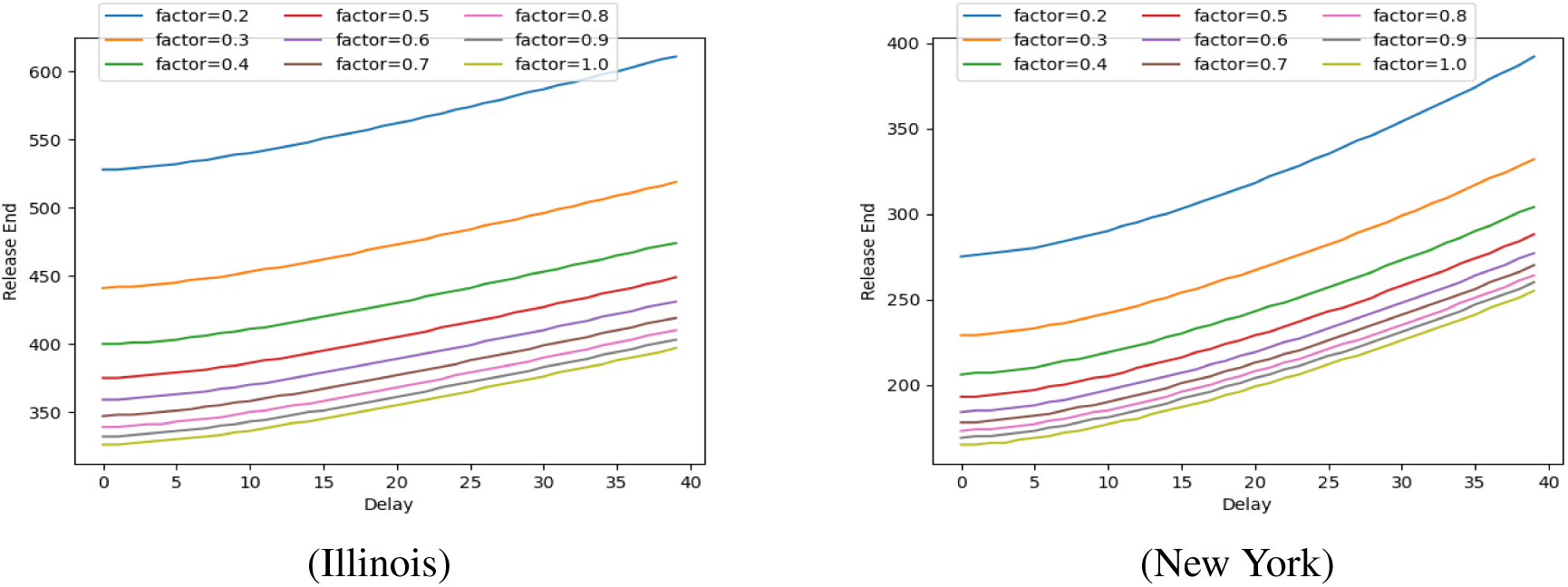
The number of days required for complete removal of lockdown vs start delay of gradual release policy for different release factors, *α_S_*.

#### SEIR Model

The SEIR model parameters were investigated and did not provide additional insight as the recovery data in the two states considered was sparse.

## 4 Model parameters

Model parameters were obtained using a program to fit the model equations. The variables used in the model are *η*, *β* and *γ*, where *η* specifies the susceptible set, i.e. *S*_0_ = *ηN*. An optimizer was used to fit determine the parameters using a loss function

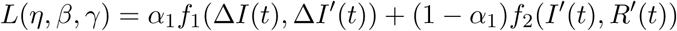

where Δ*I*′(*t*) is the derivative of the computed active infections, *I*′(*t*), *ΔI*(*t*) is the derivative of the actual data time series of active infected cases, *I*(*t*); the functions *f*_1_ was chosen to be either the RMSE, alternatively the R^2^ value (coefficient of determination) of the differences in the parameters, while *f*_2_ is again a convex combination of the RMSE, alternatively R^2^), values of differences in the two time series *I*′(*t*) and *R*′(*t*) with respect to the data time series *I*(*t*) and *R*(*t*). The data was obtained from multiple sources that include [19,20,25]. Note that the data for recovered patients does not seem to fit the average recovery time that has been reported; as such our results are to be considered with this caveat. Our model fitting results in utilizing the inverse of the reported recovery rate as a substitute for the infectious period instead of forcing that period from reports external to the model.

The results for our curve fitting are illustrated in Figure 10. We sampled over multiple dates to find the best fit, i.e., that minimizes the error function above (we in fact maximized the R^2^ value since we needed to normalize the two function *f*_1_ and *f*_2_). After some experimentation, we choose *α*_1_ = 0.3 and the convex combination weights inside *f*_2_ for *I*′(*t*) and *R*′(*t*) were 0.7 and 0.3, respectively. There is the distinct possibility and drawback of an over-fit but given the sparsity of the recovery data the determination of the degree of overfit was not conducted in this current version. The parameters obtained are *β* = 22.80, *γ* = 0.00554, *η* = 0.005724 for Illinois and *β* = 10.41, *γ* = 0.01863, *η* = 0.016788 for New York. No additional advantage is seen in the SEIR model as compared to the SIR model.

**Figure 10:**
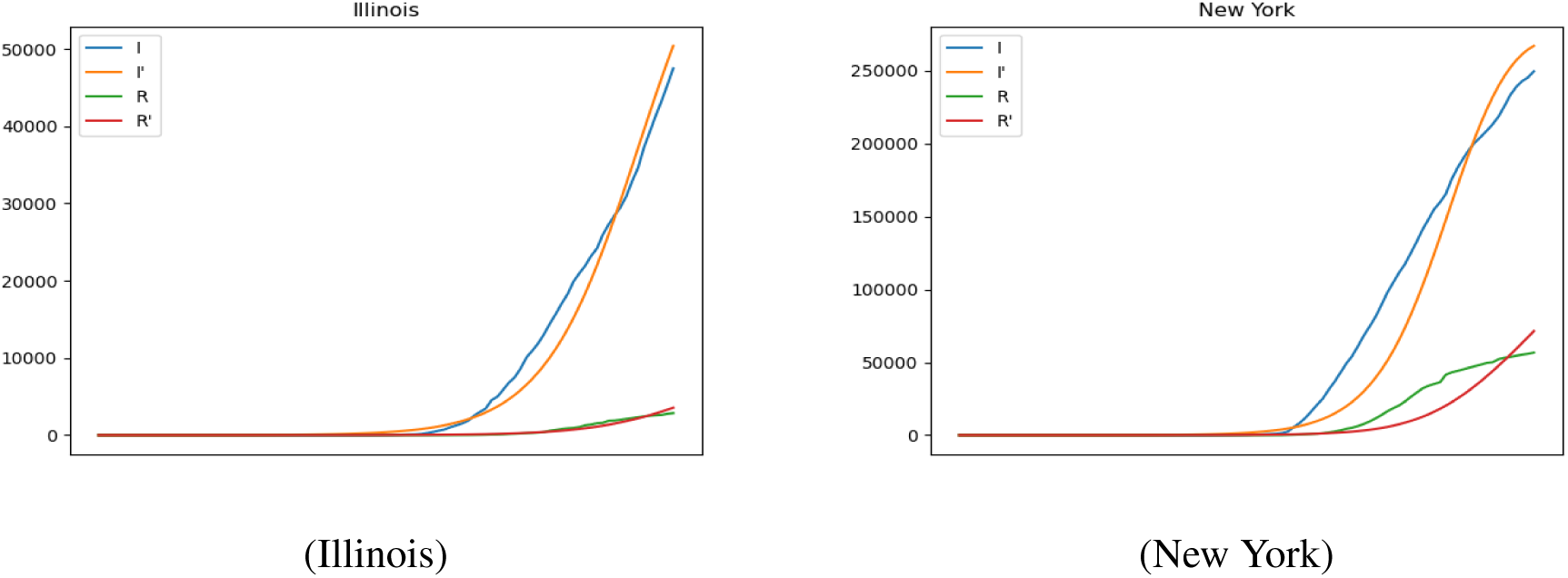
Model fitting to estimate parameters

## Data Availability

Data used is publicly available and referenced. Algorithms and implementations are available from the authors by e-mail request.

## 5 Appendix

### 5.1 Impact of adherence to lockdown in Phased release

The above relationships are all contingent to the assumption that 50% of the population were in lockdown. To illustrate the variation with the lockdown rate we consider the percentage number of days to reopen versus the lockdown factor, specified by *h* in Figure 11. As the percentage of population that is on lockdown increases, the number of days to start the first phase of the removal of the lockdown restriction increases non-linearly. This is positively countered by reduced number of infections. The threshold used in this simulation was 75%.

**Figure 11:**
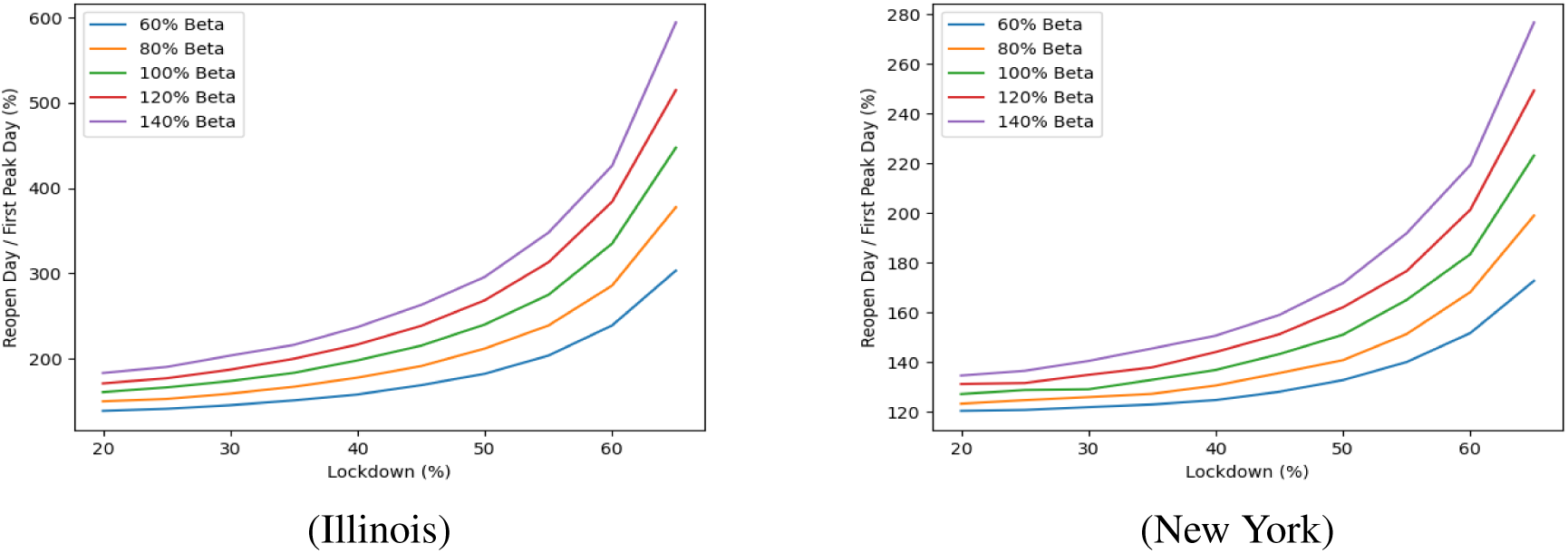
Number of days to restart / Number of days to peak vs lockdown percentage (*h*).

### 5.2 SEIR Model Parameters

Sampling over multiple dates to minimize the error function and find the best fit, we obtained the parameters *β* = 77.65839487, *β_I_* = 0.02227246, *γ* = 0.05438232, and *η* = 0.01698456 for New York, and *β* = 328.03177107, *β_I_* = 0.00525391, *γ* = 0.01430008, and *η* = 0.00625742 for Illinois. We illustrate the in Figure 12. However application of these results did not yield significant differences and will be the focus of future work.

**Figure 12.**
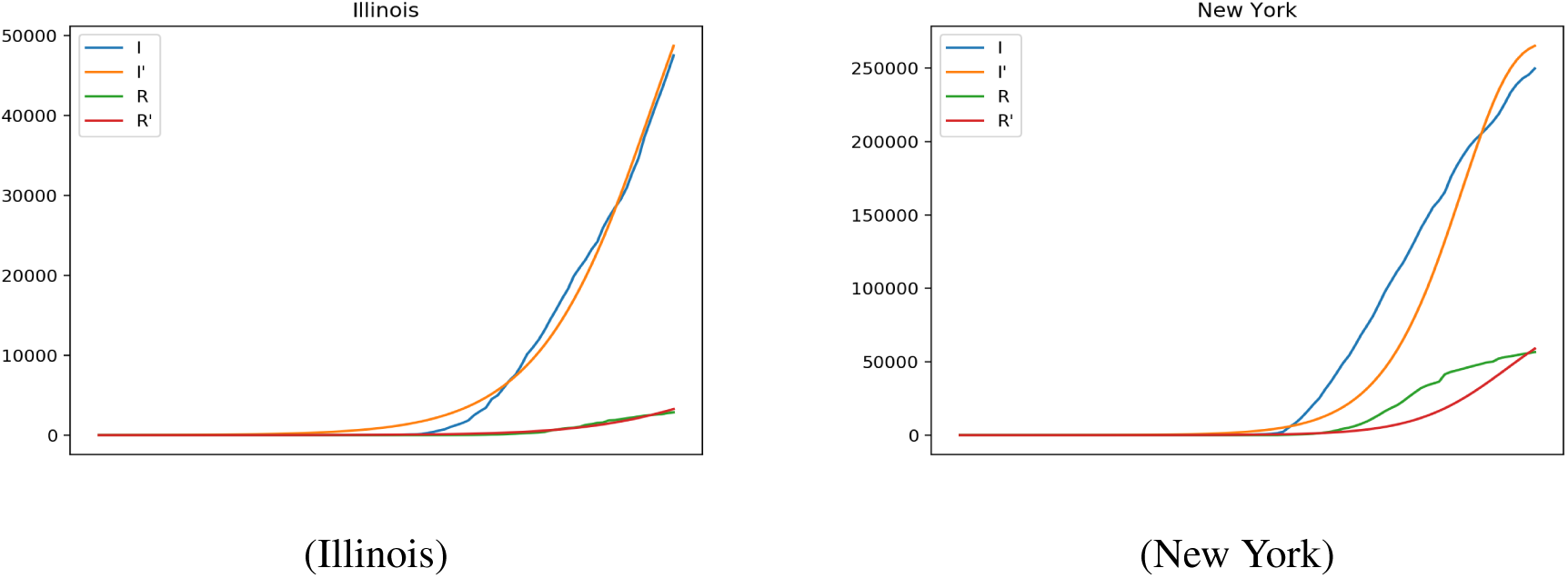
Model fitting to estimate parameters in the SEIR model results

## Footnotes

* Research sponsored by NSF grant No. 2028274. This paper may be found at http://www.cs.iit.edu/~kapoor/papers/covid1.pdf These are the results of a study at Kapoor Lab, Department of Computer Science, Illinois Tech, Chicago; conducted by Sanjiv Kapoor, Yi Zhang and Mohit Hota.

## Notes

### Competing Interest Statement

The authors have declared no competing interest.

### Funding Statement

Partially supported by NSF grant No. 2028274

